# Risk of Autoimmune Diseases Associated with Biologic Agents Targeting Type 2 Immunity: An Observational Study Using a Claims Database

**DOI:** 10.1101/2025.04.29.25326104

**Authors:** Shoichiro Inokuchi

## Abstract

**Objective.:** Biologic agents targeting type 2 immunity (anti-type 2 agents) have been implicated in the risk of autoimmune diseases. This study aimed to assess the association between anti-type 2 agents and the risk of developing autoimmune diseases.

**Methods.:** This retrospective cohort, nested case-control study utilized a large claims database in Japan. Patients diagnosed with bronchial asthma, atopic dermatitis, chronic urticaria, or eosinophilic chronic rhinosinusitis after April 2018 were included. The exposures of interest were anti-interleukin (IL)-5, anti-IL-4, and anti-IgE agents. The study outcomes included 11 autoimmune diseases and their composite. The risk of developing autoimmune diseases was assessed using two statistical models: nested case-control analysis (NCC) and time-dependent Cox proportional hazard model (tCox).

**Results.:** A total of 886,908 patients were included, with a median age of 42 years (interquartile range: 32.0–52.0). The use of the anti-IL-5 agent and anti-IgE agent showed consistent associations with the composite outcome: adjusted relative risk (RR) 2.50 (95% confidence interval [CI] 1.35, 4.64) in NCC and 3.02 (1.86, 4.90) in tCox, and 2.25 (1.37, 3.67) in NCC and 2.33 (1.61, 3.39) in tCox, respectively. Regarding individual outcomes, rheumatoid arthritis, systemic lupus erythematosus, and anti-neutrophil cytoplasmic antibody-associated vasculitis were associated with the use of anti-IL-5 agents. In patients with atopic dermatitis, anti-IL-4 agents appear to reduce the risk of psoriasis.

**Conclusion.:** Although causal interpretation is limited, it provides valuable insights into the association between anti-type 2 agents and autoimmune diseases in real-world clinical practice.

**Significance and Innovations:**

- Anti-IL-5 and anti-IgE agents are associated with an increased risk of autoimmune diseases.
- Anti-IL-4 agents show no remarkable link to autoimmune diseases but reduce psoriasis risk in atopic dermatitis.
- This study explores the effects of anti-type 2 agents and autoimmune diseases in real-world settings.

---

Type 2 immunity primarily refers to allergic immune responses characterized by the activation of type 2 immune cells, which include type 2 T helper (Th2) cells, type 2 innate lymphoid cells, eosinophils, and mast cells, along with an increase in humoral factors such as interleukin (IL)-4, IL-5, IL-9, IL-13, and IgE. Although this type of immune response is thought to have evolved as a defense mechanism against helminth infections, overactivation can lead to various conditions such as bronchial asthma (BA), atopic dermatitis (AD), chronic rhinosinusitis with nasal polyps (CRSwNP), and chronic urticaria (CU).(1,2) These diseases are generally treated with local and oral glucocorticoids, antihistamines, and immunosuppressants. In the era of biologics, several effective treatments targeting type 2 immunity have been developed including dupilumab,(3,4) which targets the IL-4 receptor; mepolizumab,(5) and benralizumab,(6) which targets IL-5 and its receptors; and omalizumab,(7) which targets IgE. These agents are generally used to treat refractory diseases.

Type 2 immunity governs not only the allergic immune response but also tissue repair and homeostasis. Therefore, blocking type 2 immunity may disrupt these regulatory mechanisms, potentially leading to harmful inflammation.(8) Agents targeting type 2 immunity have been implicated in the development of autoimmune diseases. Previous studies have reported rheumatoid arthritis (RA)-like arthritis in patients receiving omalizumab.(9,10) Regarding anti-IL-5 agents, a study in the United Kingdom found that approximately 10% of 142 BA patients experienced joint symptoms, with one patient developing RA.(11) Additionally, a case series from France documented cases of seronegative RA associated with anti-IL-5 therapy.(12) IL-4/13 inhibitors have been suggested to influence the balance between type 2 and type 1/17 immunity,(13,14) potentially triggering autoimmune diseases, such as arthritis. A registry-based study from the Netherlands(8,15) and a study using the WHO VigiBase(16) reported an association between dupilumab and the development of arthritis. Furthermore, a study conducted in the United Kingdom found that 5.8% of 400 AD patients who received dupilumab developed joint symptoms.(17) Dupilumab has also been implicated in psoriasis.(18) Additionally, a recent meta-analysis of randomized controlled trials suggested that the use of dupilumab, omalizumab, and mepolizumab in patients with CRSwNP was associated with an approximately two-fold increased risk of rheumatic adverse events compared to placebo.(19)

These findings suggest that anti-type 2 immune drugs may contribute to the development of various autoimmune conditions, including arthritis. However, no large-scale studies have comprehensively demonstrated the effects of targeting type 2 immunity. In this study, we aimed to assess the impact of biologic agents targeting type 2 immunity (anti-type 2 agents) on the development of autoimmune diseases using a large-scale claims database in Japan. The findings from this study are expected to enhance the understanding of the autoimmunity associated with type 2 immunity inhibition.

## Patients and Methods

### Data source and study design

This was a retrospective cohort, nested case-control (NCC) study conducted using a large, anonymized health insurance claims database from JMDC Inc. (Tokyo, Japan), also referred to as the JMDC Claims database.(20) The database contains the claims records of approximately 20 million individuals, covering disease diagnoses, prescriptions, and medical procedures in both inpatient and outpatient settings, as well as health checkup results. This database is widely used in various studies, including pharmacoepidemiologic studies, outcome research, and cost evaluations. The population in the database comprises individuals aged <75 years, as the health insurance societies contributing to the database do not include individuals aged ≥75 years. Since this study used a de-identified secondary database, ethical approval was not required under local regulations in Japan. The requirement for informed consent was waived, and an opt-out approach was applied in compliance with local regulations. The study was registered (ID: UMIN000057238) in the University Hospital Medical Information Network Clinical Trial Registry (Tokyo, Japan) and conducted in accordance with the principles of the Declaration of Helsinki.

### Study participants

Patients diagnosed with BA, AD, CU, or eosinophilic chronic rhinosinusitis (ECRS) between April 2018 and August 2024 were included. This cutoff was chosen based on the most recent approval date among the exposure drugs (mepolizumab, March 2016; benralizumab, April 2018; dupilumab, January 2018; omalizumab, March 2009). Definitions of AD and BA required at least two recorded diagnoses within an interval of 1–12 months, along with the use of related medications based on previous validation studies (Supplementary Table 1).(21–24) For CU and ECRS, at least two recorded diagnoses within an interval of 1–12 months were required. The first date of diagnosis was defined as the index date (day 0). Patients were then required to have continuous enrolment for ≥365 days, no prior history of the exposure drugs, be aged ≥18 years, and have a follow-up period at least 30 days. For each outcome, patients with a pre-existing diagnosis up to day 0 were excluded from the analysis.

### Exposure

Exposure was designated as an anti-IL-5 agent (mepolizumab, benralizumab), an anti-IL-4/13 agent (dupilumab), and an anti-IgE agent (omalizumab). As these biologic agents can be initiated, discontinued, or reinitiated, they were treated as time-varying exposures. Evaluation points were set at 30-day intervals in the time-dependent Cox proportional hazard (tCox) model and on the case date for cases or the date corresponding to the case date for controls in the NCC analysis. Exposure status was determined based on the presence of a prescription record within the period from 365 to 14 days before each time point, incorporating a 14-day washout period. This approach was implemented to minimize protopathic bias, where a drug administered just before the onset of an outcome may not be the cause of the outcome, but rather be prescribed in response to early clinical signs of the outcome. Detailed definitions are provided in Supplementary Table 1.

### Outcome

Study outcomes included the following autoimmune diseases and their composites: RA, systemic lupus erythematosus (SLE), systemic sclerosis (SSc), dermatomyositis/polymyositis (DM/PM), Sjögren’s syndrome (SS), mixed connective tissue disease (MCTD), anti-neutrophil cytoplasmic antibody-associated vasculitis (AAV), large-vessel vasculitis (LVV), polyarteritis nodosa (PAN), polymyalgia rheumatica (PMR), and psoriasis. SSc, DM/PM, SS, MCTD, AAV, LVV, PAN, and PMR were defined based on a validated algorithm(25) that requires two diagnostic records occurring 30–365 days apart. For RA, SLE, and psoriasis, given the relatively low accuracy of definitions based solely on diagnostic records, the definitions included two diagnostic records 30–365 days apart, accompanied by prescriptions for relevant medications.(25–30) Detailed definitions are provided in Supplementary Table 1.

### Covariates

Age, gender, baseline diseases (BA, AD, CU, and ECRS), and comorbidities included in the Charlson Comorbidity Index(31) were used as baseline covariates. Given that the outcomes involve rare diseases and the use of time-varying exposure in this study, baseline risks of each outcome were summarized by a disease risk score method,(32–34) calculated based on a Cox regression model. The partial hazard for each individual was categorized into quantiles. For time-varying variables, the prescribed medications, frequency of outpatient visits, and history of hospitalization were assessed to adjust for disease severity at each time point. Detailed covariate definitions are provided in Supplementary Table 1.

### Statistical analysis

To demonstrate the risk of exposure to outcomes while accounting for time-varying variables, the tCox model(35) and NCC analysis(36) were performed. Two different statistical models were employed to enhance the robustness of the results, considering the limitations of observational studies that utilized routinely collected health data(37). The censoring point was defined as the earliest occurrence of an outcome of interest, death, or emigration from the database. There were no missing values in this study. In the NCC analysis, non-case patients were selected at a 1:6 ratio based on the number of days from the cohort entry date (day 0) using density sampling with replacement. Patients were selected from the at-risk population who had not experienced the outcome of interest at the time of the case date (Supplementary Figure 1). Crude and covariate-adjusted relative risks (RRs) and 95% confidence intervals (CIs) of the exposures were derived using a conditional logistic regression model. The tCox models were also employed to derive crude and covariate-adjusted RRs and 95% CIs.

In addition, subgroup analyses were conducted for both the NCC and tCox models in patients with BA and AD, respectively, after adjusting for additional covariates representing disease severity (Supplementary Table 1). Sensitivity analysis was performed by extending the washout period for exposure identification from days 14 to 42. Results whose 95% CI did not include 1.0, were considered suggestive of an influence of the exposure drugs. Multiplicity was not adjusted for because of the exploratory nature of the study. Data processing was performed using Python version 3.12.3, and statistical analyses were conducted using R version 4.4.1.

## RESULTS

### Study participants

A total of 886,908 patients were included in the study, with males accounting for 46% (Figure 1). The baseline prevalence of BA, AD, ECRS, and CU were 538,908 (60.8%), 303,327 (34.2%), 3,034 (0.3%), and 48,252 (5.4%), respectively (Table 1). The median patient age was 42 years (interquartile range [IQR]: 32.0–52.0). Oral glucocorticoids were prescribed to 30,233 patients (3.4%), with a higher proportion observed in patients with ECRS (n=519, 17.1%).

**Figure 1.**
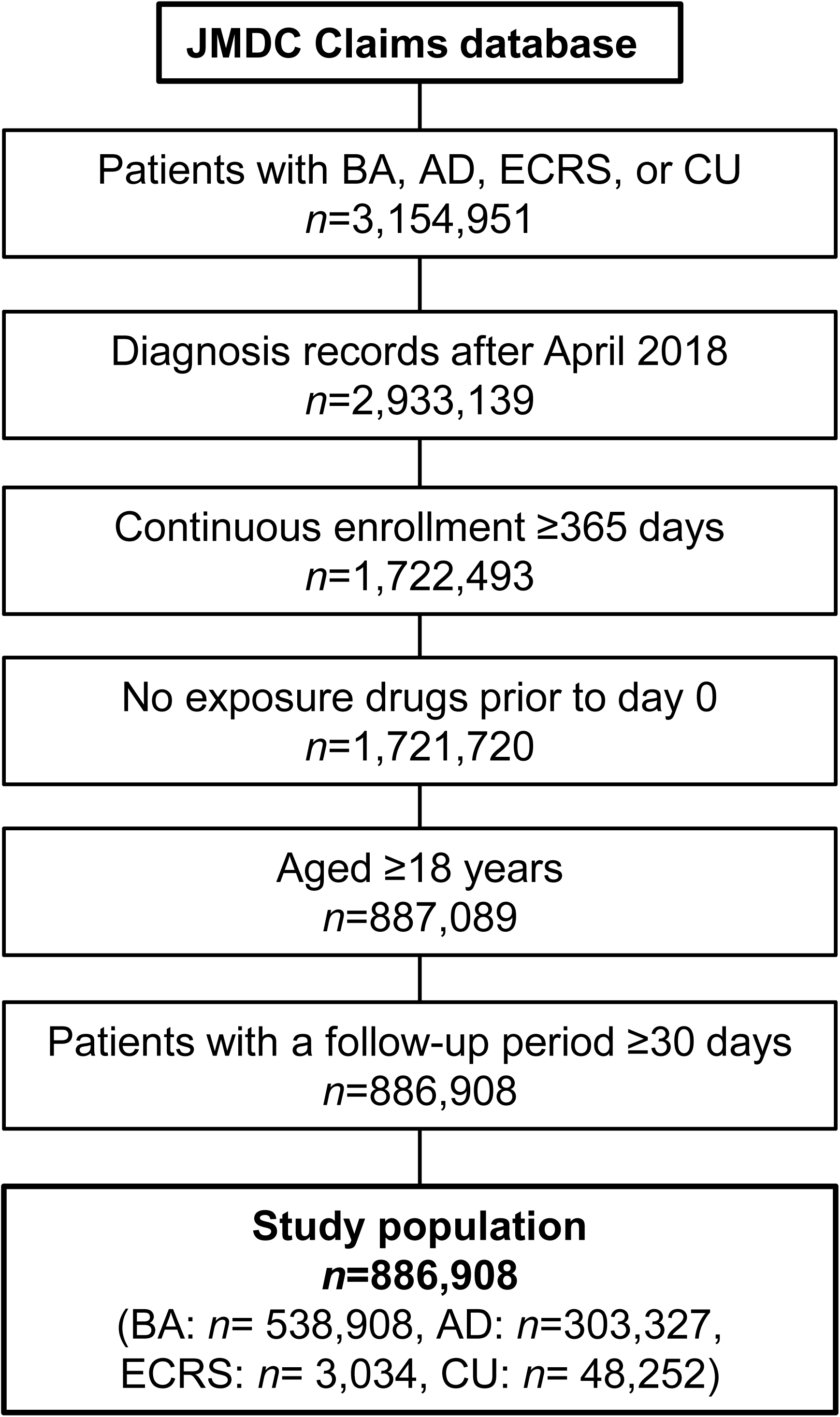
Patient flow. Abbreviations: BA, Bronchial asthma; AD, Atopic dermatitis; ECRS, Eosinophilic chronic rhinosinusitis; CU, Chronic urticaria

**Table 1.** Baseline characteristics

|  | **Total**<br>**886,908**<br>**(100.0%)** | **BA**<br>**538,908**<br>**(60.8%)** | **AD**<br>**303,327**<br>**(34.2%)** | **ECRS**<br>**3,034 (0.3%)** | **CU**<br>**48,252 (5.4%)** |
| --- | --- | --- | --- | --- | --- |
| **Demographics** |  |  |  |  |  |
| Age, median [IQR] | 42.0 [32.0, 52.0] | 44.0 [35.0, 53.0] | 37.0 [27.0, 48.0] | 49.0 [41.0, 56.0] | 46.0 [35.0, 54.0] |
| Age group |  |  |  |  |  |
| 18 ≤ 50 | 636,794 (71.8%) | 365,918 (67.9%) | 242,409 (79.9%) | 1,703 (56.1%) | 31,364 (65.0%) |
| 50 < 65 | 220,777 (24.9%) | 151,441 (28.1%) | 55,003 (18.1%) | 1,194 (39.4%) | 14,950 (31.0%) |
| 65 < 75 | 29,337 (3.3%) | 21,549 (4.0%) | 5,915 (2.0%) | 137 (4.5%) | 1,938 (4.0%) |
| Gender, male | 408,836 (46.1%) | 235,833 (43.8%) | 153,922 (50.7%) | 2,060 (67.9%) | 20,231 (41.9%) |
| **Treatment** |  |  |  |  |  |
| Oral GCs | 30,233 (3.4%) | 18,771 (3.5%) | 10,008 (3.3%) | 519 (17.1%) | 1,752 (3.6%) |
| CyA | 1,584 (0.2%) | 449 (0.1%) | 1,124 (0.4%) | 1 (0.0%) | 64 (0.1%) |
| LTRA | 88,068 (9.9%) | 72,974 (13.5%) | 13,068 (4.3%) | 1,451 (47.8%) | 2,626 (5.4%) |
| Topical GC | 129,416 (14.6%) | 28,782 (5.3%) | 96,703 (31.9%) | 123 (4.1%) | 6,526 (13.5%) |
| ICS | 16,177 (1.8%) | 14,601 (2.7%) | 1,794 (0.6%) | 126 (4.2%) | 246 (0.5%) |
| LABA | 44,324 (5.0%) | 35,349 (6.6%) | 7,974 (2.6%) | 95 (3.1%) | 1,439 (3.0%) |
| ICS/LABA | 77,820 (8.8%) | 67,617 (12.5%) | 10,322 (3.4%) | 507 (16.7%) | 1,612 (3.3%) |
| Triple therapy | 967 (0.1%) | 646 (0.1%) | 263 (0.1%) | 6 (0.2%) | 57 (0.1%) |
| Photo therapy | 14,381 (1.6%) | 5,173 (1.0%) | 8,770 (2.9%) | 19 (0.6%) | 667 (1.4%) |
| **Management** |  |  |  |  |  |
| Hospitalization history | 44,276 (5.0%) | 29,517 (5.5%) | 12,470 (4.1%) | 332 (10.9%) | 2,361 (4.9%) |
| Outpatient visit/month, median [IQR] | 0.7 [0.3, 1.3] | 0.8 [0.3, 1.3] | 0.7 [0.3, 1.2] | 1.0 [0.5, 1.6] | 0.8 [0.4, 1.4] |
| **Comorbidity** |  |  |  |  |  |
| CCI score, mean (SD) | 0.9 (1.1) | 1.1 (1.1) | 0.4 (0.9) | 0.8 (1.1) | 0.5 (1.1) |
| Follow-up days, median [IQR] | 991.0 [453.0, 1876.0] | 896.0 [421.0, 1856.0] | 1122.0 [540.0, 1938.0] | 1122.0 [540.0, 1938.0] | 1010.0 [473.0, 1765.0] |

Numbers (percentages) are shown unless otherwise specified.
Abbreviations: BA, Bronchial asthma; AD, Atopic dermatitis; ECRS Eosinophilic chronic rhinosinusitis; CCI, Charlson Comorbidity Index; CU, Chronic urticaria; CyA, Cyclosporin A; GC, Glucocorticoid; ICS, Inhaled corticosteroids; IQR, Interquartile range; LTRA, Leukotriene receptor antagonist; LABA, Long-acting $\beta$ adrenoceptor agonist.

Hospitalization history was observed in 44,276 patients (5.0%), while the median number of outpatient visits was 0.7 times/month (IQR: 0.3–1.3). The median follow-up period in the study cohort was 991.0 days (IQR: 453.0–1876.0).

### Impact of anti-IL-5 agents on autoimmune diseases

In the composite outcome, the NCC analysis yielded a crude RR of 4.62 (95% CI: 2.58, 8.27) and an adjusted RR of 2.50 (1.35, 4.64), whereas the tCox analysis resulted in RR of 5.18 (3.22, 8.35) and 3.02 (1.86, 4.90), respectively, indicating an increased risk associated with anti-IL-5 agents. When examining each outcome individually, positive estimates that did not cross 1.0 were observed in RA (NCC adjusted: 4.98 [1.85, 13.37]; tCox adjusted: 5.03 [2.81, 9.00]), SLE (NCC adjusted: 15.49 [2.32, >100]; tCox adjusted: 12.40 [5.62, 27.33]), and AAV (NCC adjusted: 42.18 [4.18, >100]; tCox adjusted: 14.71 [7.45, 29.04]) (Table 2). Among AAV cases, eosinophilic granulomatosis with polyangiitis (EGPA) was the most prevalent, especially in patients treated with anti-IL-5 agents (Supplementary Table 2). In the BA subgroup, a trend similar to that observed in the overall population was noted.The risk of psoriasis was elevated only in the crude analysis, with 3.00 (1.03, 8.78) for NCC and 2.58 (1.07, 6.20) for tCox (Supplementary Table 3). A limited number of patients receiving anti-IL-5 agents and experiencing the outcomes were observed in the AD subgroup (Supplementary Table 4). The sensitivity analysis, in which the washout period used to define exposure was extended to 42 days to ensure robustness, yielded similar findings, including a strong association between anti-IL-5 agents and AAV was observed (NCC adjusted: 18.90 [2.14, >100]; tCox adjusted: 12.20 [5.82, 25.55], Supplementary Table 5).

**Table 2.** Effect of anti-IL-5 agents on the outcomes

| Disease | NCC |  |  |  | tCox |  |  |  |
| --- | --- | --- | --- | --- | --- | --- | --- | --- |
|  | Case<br>Ex/N | Control<br>Ex/N | Crude | Adjusted | Ex<br>Ev/PY | Control<br>Ev/PY | Crude | Adjusted |
| **Composite** | 20/11,989 | 26/71,934 | **4.62**<br>[2.58, 8.27] | **2.50**<br>[1.35, 4.64] | 17/850.9 | 11,972/2,662,492.2 | **5.18**<br>[3.22, 8.35] | **3.02**<br>[1.86, 4.90] |
| **RA** | 12/2,195 | 10/13,170 | **7.20**<br>[3.11, 16.67] | **4.98**<br>[1.85, 13.37] | 12/1,372.1 | 2,183/2,747,401.3 | **11.74**<br>[6.65, 20.71] | **5.03**<br>[2.81, 9.00] |
| **SLE** | 8/602 | 3/3612 | **22.43**<br>[4.74, 100<] | **15.49**<br>[2.32, 100<] | 7/1,352.9 | 595/2,763,328.4 | **29.12**<br>[13.63, 62.22] | **12.40**<br>[5.62, 27.33] |
| **SSc** | 1/379 | 1/2,274 | 6.00<br>[0.38, 95.93] | 5.64<br>[0.28, 100<] | 1/1,440.7 | 378/2,767,550.1 | 5.84<br>[0.82, 41.66] | 2.57<br>[0.35, 18.86] |
| **DM/PM** | 1/653 | 4/3,918 | 1.50<br>[0.17, 13.42] | 0.50<br>[0.05, 4.59] | 1/1,425.6 | 652/2,766,616.9 | 3.31<br>[0.46, 23.63] | 1.61<br>[0.22, 11.48] |
| **SS** | 2/1,950 | 8/11,700 | 1.50<br>[0.32, 7.06] | 1.09<br>[0.19, 6.24] | 2/1,396.6 | 1,948/2,751,632.0 | 2.24<br>[0.56, 8.97] | 0.98<br>[0.25, 3.90] |
| **MCTD** | 0/143 | 0/858 | NA<br>[NA, NA] | NA<br>[NA, NA] | 0/1,450.3 | 143/2,769,033.1 | **0.00**<br>[0.00, 0.00] | **0.00**<br>[0.00, 0.00] |
| **AAV** | 14/748 | 1/4,488 | **84.00**<br>[11.05, 100<] | **42.18**<br>[4.18, 100<] | 11/947.7 | 737/2,766,649.7 | **47.61**<br>[25.95, 87.36] | **14.71**<br>[7.45, 29.04] |
| **LVV** | 0/75 | 0/450 | NA<br>[NA, NA] | NA<br>[NA, NA] | 0/1,447.8 | 75/2,770,357.5 | **0.00**<br>[0.00, 0.00] | **0.00**<br>[0.00, 0.00] |
| **PAN** | 1/51 | 0/306 | 100<<br>[0.00, 100<] | 100<<br>[0.00, 100<] | 1/1,429.1 | 50/2,770,496.9 | **44.32**<br>[6.00, 100<] | **23.72**<br>[3.02, 100<] |
| **PMR** | 1/325 | 4/1,950 | 1.50<br>[0.17, 13.42] | 0.34<br>[0.03, 4.25] | 1/1,443.9 | 324/2,769,177.6 | 6.39<br>[0.90, 45.61] | 2.49<br>[0.35, 17.68] |
| **Psoriasis** | 6/7214 | 22/43,284 | 1.64<br>[0.66, 4.04] | 1.45<br>[0.55, 3.86] | 5/1,415.1 | 7,209/2,711,247.3 | 1.64<br>[0.68, 3.93] | 1.51<br>[0.62, 3.63] |

The effect of the exposure drug is presented as the odds ratio [95% CI] for the nested case-control analysis and the hazard ratio [95% CI] for the time-dependent Cox analysis.
Results with a 95% CI that does not include 1.0 are indicated in bold.
Abbreviations: Ex, Exposure; Ev, Event; NA, Not applicable; NCC, Nested case-control analysis; PY, Patient-year; tCox, Time-dependent Cox analysis.

### Impact of anti-IL-4/13 agents on autoimmune diseases

The composite outcome showed a crude RR of 1.36 (95% CI: 1.01, 1.83) for NCC and 1.52 (1.14, 2.02) for tCox, while the adjusted RRs were 0.86 (0.63, 1.17) and 1.02 (0.77, 1.35), respectively (Table 3). Similar findings were observed for AAV and psoriasis, with crude RRs of 3.50 (1.38, 8.89) in NCC and 2.60 (1.16, 5.83) in tCox for AAV, and 1.43 (1.00, 2.04) in NCC and 1.83 (1.29, 2.57) in tCox for psoriasis, without any remarkable association observed in the adjusted models. In the BA subgroup analysis, these findings were largely consistent with those observed in the overall population (Supplementary Table 3). The AD subgroup analysis demonstrated a consistent protective effect of anti-IL-4/13 agents on the composite outcome, with adjusted RRs of 0.50 (0.34, 0.74) for NCC and 0.64 (0.45, 0.91) for tCox. The analysis of individual outcomes indicated that the effect on psoriasis was particularly pronounced, with adjusted RRs of 0.54 (0.35, 0.85) for NCC and 0.68 (0.47, 1.00) for tCox (Supplementary Table 4). The sensitivity analysis, in which the washout period was extended to 42 days, demonstrated a trend similar to that observed in the main analysis (Supplementary Table 5).

**Table 3.** Effect of anti-IL-4/13 agents on the outcomes

| Disease | NCC |  |  |  | tCox |  |  |  |
| --- | --- | --- | --- | --- | --- | --- | --- | --- |
|  | Case<br>Ex/N | Control<br>Ex/N | Crude | Adjusted | Ex<br>Ev/PY | Control<br>Ev/PY | Crude | Adjusted |
| **Composite** | 54/11,989 | 239/71,934 | **1.36**<br>**[1.01, 1.83]** | 0.86<br>[0.63, 1.17] | 48/8,325.0 | 11941/2,655,018.1 | **1.52**<br>**[1.14, 2.02]** | 1.02<br>[0.77, 1.35] |
| **RA** | 10/2,195 | 57/13,170 | 1.05<br>[0.54, 2.07] | 1.06<br>[0.46, 2.40] | 9/9,274.7 | 2186/2,739,498.7 | 1.31<br>[0.68, 2.52] | 1.24<br>[0.64, 2.40] |
| **SLE** | 1/602 | 13/3,612 | 0.46<br>[0.06, 3.53] | 0.56<br>[0.06, 5.34] | 1/9,307.7 | 601/2,755,373.6 | 0.60<br>[0.08, 4.27] | 0.41<br>[0.06, 3.00] |
| **SSc** | 1/379 | 10/2,274 | 0.60<br>[0.08, 4.69] | 0.72<br>[0.03, 18.17] | 1/9,310.4 | 378/2,759,680.4 | 0.91<br>[0.13, 6.50] | 0.90<br>[0.12, 6.51] |
| **DM/PM** | 0/653 | 18/3,918 | 0.00<br>[0.00, 100<] | 0.00<br>[0.00, 100<] | 0/9,302.4 | 653/2,758,740.1 | **0.00**<br>**[0.00, 0.00]** | **0.00**<br>**[0.00, 0.00]** |
| **SS** | 10/1,950 | 51/11,700 | 1.18<br>[0.60, 2.32] | 1.17<br>[0.53, 2.56] | 10/9,198.1 | 1940/2,743,830.5 | 1.73<br>[0.93, 3.22] | 1.70<br>[0.91, 3.16] |
| **MCTD** | 0/143 | 7/858 | 0.00<br>[0.00, 100<] | 0.00<br>[0.00, 100<] | 0/9,314.5 | 143/2,761,168.8 | **0.00**<br>**[0.00, 0.00]** | **0.00**<br>**[0.00, 0.00]** |
| **AAV** | 7/748 | 12/4,488 | **3.50**<br>**[1.38, 8.89]** | 2.00<br>[0.55, 7.31] | 6/9,269.1 | 742/2,758,328.3 | **2.60**<br>**[1.16, 5.83]** | 1.98<br>[0.85, 4.60] |
| **LVV** | 1/75 | 3/450 | 2.00<br>[0.21, 19.23] | 1.35<br>[0.11, 16.86] | 1/9,317.9 | 74/2,762,487.4 | 4.36<br>[0.58, 32.55] | 4.06<br>[0.54, 30.36] |
| **PAN** | 0/51 | 1/306 | 0.00<br>[0.00, 100<] | 0.00<br>[0.00, 100<] | 0/9,319.0 | 51/2,762,607.0 | **0.00**<br>**[0.00, 0.00]** | **0.00**<br>**[0.00, 0.00]** |
| **PMR** | 0/325 | 6/1950 | 0.00<br>[0.00, 100<] | 0.00<br>[0.00, 100<] | 0/9,310.0 | 325/2,761,311.5 | **0.00**<br>**[0.00, 0.00]** | **0.00**<br>**[0.00, 0.00]** |
| **Psoriasis** | 37/7,214 | 156/43284 | 1.43<br>[1.00, 2.04] | **0.58**<br>**[0.40, 0.85]** | 33/8,515.1 | 7181/2,704,147.3 | **1.83**<br>**[1.29, 2.57]** | 0.97<br>[0.69, 1.37] |

The effect of the exposure drug is presented as the odds ratio [95% CI] for the nested case-control analysis and the hazard ratio [95% CI] for the time-dependent Cox analysis.
Results with a 95% CI that does not include 1.0 are indicated in bold.
Abbreviations: Ex, Exposure; Ev, Event; NA, Not applicable; NCC, Nested case-control analysis; PY, Patient-year; tCox, Time-dependent Cox analysis.

### Impact of anti-IgE agents on autoimmune diseases

For the composite outcome, the NCC analysis yielded a crude RR of 3.79 (95% CI: 2.39, 5.99) and an adjusted RR of 2.25 (1.37, 3.67), while the tCox analysis resulted in RRs of 3.31 (2.28, 4.79) and 2.33 (1.61, 3.39), respectively, indicating an increased risk of autoimmune diseases associated with anti-IgE agents (Table 4). When examining individual outcomes, positive estimates that did not cross 1.0 were observed for psoriasis, with adjusted RRs of 3.43 (1.73, 6.80) for NCC and 2.56 (1.59, 4.13) for tCox. For SS and AAV, an increased risk was observed in the crude analysis (4.80 [1.29, 17.88] and 24.00 [2.68, 100<] for NCC, and 2.76 [1.03, 7.35] and 7.23 [2.71, 19.33] for tCox).

**Table 4.**
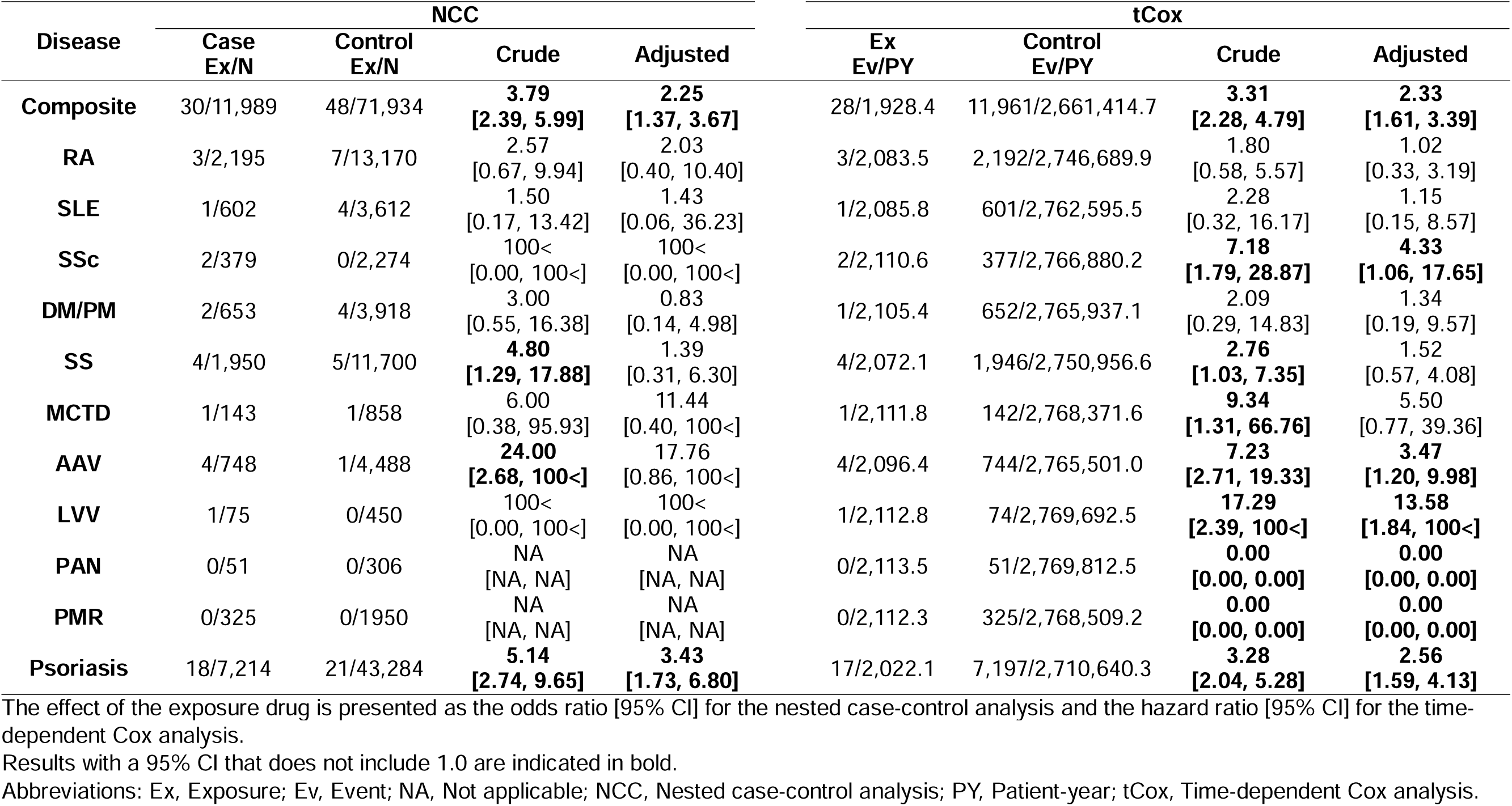
Effect of anti-IgE agents on the outcomes

| Disease | NCC |  |  |  | tCox |  |  |  |
| --- | --- | --- | --- | --- | --- | --- | --- | --- |
|  | Case<br>Ex/N | Control<br>Ex/N | Crude | Adjusted | Ex<br>Ev/PY | Control<br>Ev/PY | Crude | Adjusted |
| **Composite** | 30/11,989 | 48/71,934 | **3.79**<br>**[2.39, 5.99]** | **2.25**<br>**[1.37, 3.67]** | 28/1,928.4 | 11,961/2,661,414.7 | **3.31**<br>**[2.28, 4.79]** | **2.33**<br>**[1.61, 3.39]** |
| **RA** | 3/2,195 | 7/13,170 | 2.57<br>[0.67, 9.94] | 2.03<br>[0.40, 10.40] | 3/2,083.5 | 2,192/2,746,689.9 | 1.80<br>[0.58, 5.57] | 1.02<br>[0.33, 3.19] |
| **SLE** | 1/602 | 4/3,612 | 1.50<br>[0.17, 13.42] | 1.43<br>[0.06, 36.23] | 1/2,085.8 | 601/2,762,595.5 | 2.28<br>[0.32, 16.17] | 1.15<br>[0.15, 8.57] |
| **SSc** | 2/379 | 0/2,274 | 100<<br>[0.00, 100<] | 100<<br>[0.00, 100<] | 2/2,110.6 | 377/2,766,880.2 | **7.18**<br>**[1.79, 28.87]** | **4.33**<br>**[1.06, 17.65]** |
| **DM/PM** | 2/653 | 4/3,918 | 3.00<br>[0.55, 16.38] | 0.83<br>[0.14, 4.98] | 1/2,105.4 | 652/2,765,937.1 | 2.09<br>[0.29, 14.83] | 1.34<br>[0.19, 9.57] |
| **SS** | 4/1,950 | 5/11,700 | **4.80**<br>**[1.29, 17.88]** | 1.39<br>[0.31, 6.30] | 4/2,072.1 | 1,946/2,750,956.6 | **2.76**<br>**[1.03, 7.35]** | 1.52<br>[0.57, 4.08] |
| **MCTD** | 1/143 | 1/858 | 6.00<br>[0.38, 95.93] | 11.44<br>[0.40, 100<] | 1/2,111.8 | 142/2,768,371.6 | **9.34**<br>**[1.31, 66.76]** | 5.50<br>[0.77, 39.36] |
| **AAV** | 4/748 | 1/4,488 | **24.00**<br>**[2.68, 100<]** | 17.76<br>[0.86, 100<] | 4/2,096.4 | 744/2,765,501.0 | **7.23**<br>**[2.71, 19.33]** | **3.47**<br>**[1.20, 9.98]** |
| **LVV** | 1/75 | 0/450 | 100<<br>[0.00, 100<] | 100<<br>[0.00, 100<] | 1/2,112.8 | 74/2,769,692.5 | **17.29**<br>**[2.39, 100<]** | **13.58**<br>**[1.84, 100<]** |
| **PAN** | 0/51 | 0/306 | NA<br>[NA, NA] | NA<br>[NA, NA] | 0/2,113.5 | 51/2,769,812.5 | **0.00**<br>**[0.00, 0.00]** | **0.00**<br>**[0.00, 0.00]** |
| **PMR** | 0/325 | 0/1950 | NA<br>[NA, NA] | NA<br>[NA, NA] | 0/2,112.3 | 325/2,768,509.2 | **0.00**<br>**[0.00, 0.00]** | **0.00**<br>**[0.00, 0.00]** |
| **Psoriasis** | 18/7,214 | 21/43,284 | **5.14**<br>**[2.74, 9.65]** | **3.43**<br>**[1.73, 6.80]** | 17/2,022.1 | 7,197/2,710,640.3 | **3.28**<br>**[2.04, 5.28]** | **2.56**<br>**[1.59, 4.13]** |

The effect of the exposure drug is presented as the odds ratio [95% CI] for the nested case-control analysis and the hazard ratio [95% CI] for the time-dependent Cox analysis.
Results with a 95% CI that does not include 1.0 are indicated in bold.
Abbreviations: Ex, Exposure; Ev, Event; NA, Not applicable; NCC, Nested case-control analysis; PY, Patient-year; tCox, Time-dependent Cox analysis.

Additionally, although the results between NCC and tCox were inconsistent for SSc and LVV due to the absence of outcomes in the control group in the NCC analysis, strong associations were observed in the tCox model, with adjusted RRs of 4.33 (1.06, 17.65) and 13.58 (1.84, 100<), respectively. In the BA subgroup, an increased risk for the composite outcome was observed in the crude analysis: 5.33 [2.06, 13.82] for NCC and 2.73 [1.31, 5.70] for tCox (Supplementary Table 3). In the AD subgroup, increased risks were suggested in the crude analysis for both the composite outcome and psoriasis, with crude RRs of 6.00 (2.38, 15.12) and 4.50 (1.56, 12.97) for NCC, and 4.75 (2.47, 9.14) and 3.86 (1.74, 8.54) for tCox, respectively (Supplementary Table 4). The sensitivity analysis, in which the washout period was extended to 42 days, showed that the direction of the estimates was consistent with the main analysis, supporting the robustness of the findings (Supplementary Table 5).

## DISCUSSION

In this study, we investigated the association between anti-type 2 agents and the development of autoimmune diseases. To the best of our knowledge, this is the first observational study to examine the risks associated with anti-type 2 agents using a large sample size. Although it was an observational study using a claims database and subject to limitations such as residual confounding,(37) the findings indicated an increased risk of several autoimmune diseases in patients receiving these biologic agents.

Previous studies have suggested that anti-IL-5 agents may induce autoimmune diseases, including arthritis.(11,12) Consistent with these reports, our study demonstrated an increased risk of autoimmune diseases, including RA. Furthermore, our findings indicate that SLE and AAV may be significantly associated with anti-IL-5 use. For AAV, the presence of subclinical EGPA before the initiation of anti-IL-5 therapy should be considered. Patients with EGPA often exhibit severe manifestations of underlying conditions, such as asthma and rhinosinusitis, before disease onset. These symptoms may have led to the administration of anti-IL-5 agents, potentially contributing to the observed risk. Additionally, reverse causation (i.e., the diagnosis of EGPA leading to the prescription of anti-IL-5 agents) may explain this association. However, the sensitivity analysis extending the wash-out period for exposure identification to 42 days suggested that the effect of reverse causation was only partial. The use of anti-IL-5 agents may increase the risk of autoimmune diseases in real-world clinical practice, although causal links require further study.

IL-5 is primarily involved in the proliferation, maturation, and activation of eosinophils.(8) Although eosinophils play a role in anti-helminth immunity and tissue damage,(38) they also contribute to tissue homeostasis and suppress excessive immune responses.(39) Additionally, eosinophil depletion disrupts the balance between Th17 and regulatory T cells (Tregs).(40,41) Anti-IL-5 agents may contribute to the development of autoimmune diseases by depleting eosinophils, which play homeostatic roles.

In the analysis of anti-IL-4/13 agents, an increased risk was observed in the crude analysis of the composite outcome, AAV, and psoriasis. However, after adjusting for covariates, no clear association was observed, suggesting that the severity of type 2 immune diseases may contribute to the risk of autoimmune diseases. In the AD subgroup, anti-IL-4/13 agents appeared to be associated with a reduced risk of autoimmune diseases, particularly psoriasis. This finding contradicts previous reports suggesting the development of psoriasis after anti-IL-4/13 therapy in AD patients.(18) Subgroup analyses of BA and AD showed that the observed risk of psoriasis was nearly three times higher in AD than in BA (BA: 2,312/1,607,396.1 = 0.14/100 patient-years; AD: 4,639/968,737.4 = 0.48/100 patient-years; Supplementary Tables 3 and 4), suggesting that this difference may have influenced previous findings regarding the occurrence of psoriasis in AD patients.

IL-4 is also a key cytokine involved in type 2 immunity; however, its role and mechanism of action differ from those of IL-5. IL-4 has diverse functions, including differentiation of Th2 cells, proliferation and class switching of B cells, and induction of alternatively activated macrophages, which facilitate tissue regulation and fibrosis.(42) In SLE, IL-4 produced by type 2 follicular helper T cells has been suggested to contribute to autoantibody production in SLE.(43) Although the 95% CIs for the estimated RR of the anti-IL-4/13 agent included 1.0, the point estimates for both NCC and tCox were approximately 0.5, indicating a potential pathogenic role of IL-4 in SLE. The estimated risk of psoriasis was reduced in AD patients treated with anti-IL-4/13 agents, which contradicts from the previous study.(18) A subset of AD patients exhibits both Th2 and Th17 signatures, with an intermediate gene expression pattern between those with a pure Th2 phenotype and psoriasis.(44) Furthermore, activation of both Th2 and Th17 signatures has been observed in psoriasis associated with dupilumab use, and treatment with dupilumab has been shown to modulate not only Th2 signatures but also Th17-related molecules and proinflammatory cytokines.(18) These findings suggest that dupilumab may regulate the development of psoriasis in AD by modulating both Th2 and Th17 immune responses. However, given the inconsistencies with previous reports, further research is required to clarify the relationship between IL-4/13 inhibition and psoriasis development.

In this study, an increased risk of the composite outcome was observed for anti-IgE agents. Among the diseases examined, psoriasis showed consistent results in both NCC and tCox analyses, whereas diseases such as SSc, SS, and AAV also demonstrated numerically high estimated RRs. Previous reports have indicated the development of arthritis following omalizumab treatment.(9,10) The RA results did not demonstrate a significant association with anti-IgE agent use; however, the crude point estimates for both NCC and tCox were approximately 2.0. Although a potential therapeutic effect of omalizumab has been suggested,(45) our findings did not indicate any protective effects of omalizumab against SLE development.

While no clear reports exist on the specific mechanisms by which omalizumab may lead to autoimmune diseases, it has been shown to increase the expression of various cytokines, including IL-2, IL-5, IL-6, IL-22, and TNF-α, following treatment.(46) It has also been reported to enhance antiviral responses in plasmacytoid dendritic cells by promoting type I interferon responses.(47) These findings may partially explain the development of autoimmune diseases, including psoriasis, in patients treated with omalizumab. In contrast, omalizumab has been reported to contribute to Treg induction.(48) Further investigation is required to elucidate the underlying mechanisms.

This observational study examined the association between anti-type 2 agents and the development of autoimmune diseases using a large sample size, which is a notable strength. However, it is important to acknowledge several limitations that are primarily inherent to the use of a routinely collected database. First, owing to insurance coverage, patients aged ≥75 years were not included in the database used in this study. The results may be limited regarding diseases with a higher incidence in older populations, such as PMR. Second, although the study identified patients with autoimmune disease based on well-validated methods in previous studies, an accurate judgement of disease onset requires a more detailed physical examination and meticulous chart reviews, potentially introducing a misclassification bias. Third, the database did not contain laboratory test results, making it impossible to perform a detailed assessment of the affected patients.

Fourth, despite the large sample size, the frequency of anti-type 2 agents and incidence of autoimmune diseases were low, resulting in wide CIs for some outcomes and limiting the precision of the study. Therefore, further validation using other data sources is required. Finally, as the database is primarily derived from claims data, it does not account for potential confounders, such as physical examination, laboratory test results, and pathological heterogeneity. Given the possibility of residual confounding, the results should be interpreted as descriptive insights from real-world clinical practice rather than as causal relationships.

In conclusion, we investigated the association between anti-type 2 agents and the development of autoimmune diseases using a claims database and observed an increased risk of several autoimmune diseases, especially for anti-IL-5 and anti-IgE agents. In contrast to previous reports, the findings suggest a potentially protective effect of anti-IL-4/13 against psoriasis among patients with AD. Given the limitations inherent to databases derived from claims data, further research is required to establish causal associations. Nevertheless, this study provides valuable insights into the association between type 2 immunity and autoimmune diseases.

## Data Availability

The data utilized for this research will be made available by JMDC Inc. upon reasonable request to the corresponding author.

## ACKNOWLEDGMENTS

The author thanks Mr. Takumi Tajima and Mr. Gen Terashima from JMDC Inc. (Tokyo, Japan) for their contribution to the resources, funding acquisition, and supervision of the study. The author also thanks Dr. Satomi Inokuchi from Fukuoka Heartnet Hospital (Fukuoka, Japan) for the valuable discussions. During the preparation of this work the author(s) used ChatGPT (openai.com/index/chatgpt, accessed on 22 March 2025) and Paperpal (paperpal.com, accessed on 22 March 2025) for English editing in some sentences. After using this tool/service, the author(s) reviewed and edited the content as needed and take(s) full responsibility for the content of the publication. The author thanks Editage (www.editage.jp) for the English language editing.

