## Supplementary file for "Risk of Autoimmune Diseases Associated with Biologic Agents Targeting Type 2 Immunity: An Observational Study Using a Claims Database"

**Supplementary materials**

| **Supplementary Table 1. Variable definition** | | | | |
| --- | --- | --- | --- | --- |
| **Variable** | | | | **Definition** |
| **Study participant definition** | | | | |
|  | | Bronchial asthma | | ICD-10 code: J45 |
|  | | Related medications^†^ | | (WHO-ATC code) - Inhaled corticosteroids (ICS): R03BA ,R03AK06, R03AK07, R03AK10, R03AK11, R03AK14, R03AL08, R03AL12 - Long-acting beta-agonists (LABA): R03AC12, R03AC13, R03AC18, R03AK06, R03AK07, R03AK10, R03AK11, R03AK14, R03AL08, R03AL12, R03CC11 (External use only), R03CC13 - Leukotriene receptor antagonist: R03DC01, R03DC02, R03DC03 - Xanthine: R03DA - Systemic glucocorticoids: H02AB (Oral use only) |
|  | | Atopic dermatitis | | ICD-10 code: L20 |
|  | | Related medications^†^ | | (WHO-ATC code) Topical glucocorticoids: D05AX52, D07A, D07CB01, D07CC, D07XA01 (7-digit drug code^‡^: 2649861, 2649800), D07XB05 (7-digit drug code^‡^: 2649854, 2649801) |
|  | | Chronic urticaria | | ICD-10 code (Standard disease code): L501 (8841381), L508 (8841393, 7088008) |
|  | | Eosinophilic chronic rhinosinusitis | | ICD-10 code: J328 |
| **Exposure definitions** | | | |  |
|  | | Mepolizumab | | WHO-ATC code: R03DX09 |
|  | | Benralizumab | | WHO-ATC code: R03DX10 |
|  | | Dupilumab | | WHO-ATC code: D11AH05 |
|  | | Omalizumab | | WHO-ATC code: R03DX05 |
| **Outcome definition** | | | |  |
|  | | Rheumatoid arthritis | | ICD-10 code (Standard disease code): M05, M0601–M0609, M068, M069, M0600 (8842105, 8833113) |
|  | | Related medications^†^ | | (WHO-ATC code) - Tumor necrosis alpha inhibitors: L04AB - Interleukin-6 inhibitors: L04AC07, L04AC14 - Janus kinase inhibitors: L04AF01, L04AF02, L04AF03, L04AF04, L04AF06 - Abatacept: L04AA24 - Methotrexate: L01BA01, L04AX03 - Sulfasalazine: A07EC01 - Tacrolimus: L04AD02 - Leflunomide: L04AK01 - Mizoribine: L04AX (7-digit drug code^‡^: 3999002) - Other csDMARDs: M01C |
|  | | Systemic lupus erythematosus | | ICD-10 code: M32 |
|  | | Related medications^†^ | | (WHO-ATC code) - Systemic glucocorticoids: H02AB (Oral use only) |
|  | | Psoriasis | | ICD-10 code: L40 |
|  | | Related medications^†^ | | (WHO-ATC code) - Topical vitamin D: D05AX02, D05AX04, D05AX52, D05AX (7-digit drug code^‡^: 2691702, 2699804) - Topical glucocorticoids: D05AX52, D07A, D07CB01, D07CC, D07XA01 (7-digit drug code^‡^: 2649861, 2649800), D07XB05 (7-digit drug code^‡^: 2649854, 2649801) - Tildrakizumab: L04AC17 |
|  | | ANCA-associated vasculitis | | ICD-10 code (Standard disease code): M301, M313, M317, M318 (8845513) |
|  | | Dermatomyositis/Polymyositis | | ICD-10 code: M33, M360 |
|  | | Large vessel vasculitis | | ICD-10 code: M314–M316 |
|  | | Mixed connective tissue disease | | ICD-10 code (Standard disease code): M351 (7109008, 7109007) |
|  | | Polyarteritis nodosa | | ICD-10 code: M300, M302, M308 |
|  | | Polymyalgia rheumatica | | M353 |
|  | | Sjogren syndrome | | ICD-10 code: M350 |
|  | | Systemic sclerosis | | ICD-10 code: M34 |
| **Covariates** | | | |  |
|  | Disease risk score | | The individual disease risk score was derived from a Cox proportional hazard model for each outcome, developed using the total study population. The variables considered were age (using cubic spline interpolation), gender, bronchial asthma, atopic dermatitis, chronic urticaria, eosinophilic chronic rhinosinusitis, and each of the 17 components of the Charlson Comorbidity Index at baseline. Partial hazard estimates for individual patients were categorized into quantiles and incorporated into the outcome model. | |
|  | History of hospitalisation | | Hospitalisation history from 365 to 90 days before each time point. | |
|  | Frequency of outpatient visits per month | | The number of outpatient visits recorded between 365 and 90 days before each time point was divided by 276 and then multiplied by 30. The logarithm of this value, after adding 0.5, was incorporated into the model. | |
|  | | **[Components in Charlson Comorbidity Index (ICD-10 code)]** * The following diseases are defined based on records from day -365 to day 0 | | |
|  | | Myocardial infarction | | I21, I22, I252 |
|  | | Congestive heart failure | | I099, I110, I130, I132, I255, I420, I425–I429, I43, I50, P290 |
|  | | Peripheral vascular disease | | I70, I71, I731, I738, I739, I771, I790, I792, K551, K558, K559, Z958, Z959 |
|  | | Cerebrovascular disease | | G45, G46, H340, I60–I69 |
|  | | Dementia | | F00–F03, F051, G30, G311 |
|  | | Chronic pulmonary disease | | I278, I279, J40–J47, J60–J67, J684, J701, J703 |
|  | | Rheumatic disease | | M05, M06, M315, M32–M34, M351, M353, M360 |
|  | | Peptic ulcer disease | | K25–K28 |
|  | | Mild liver disease | | B18, K700–K703, K709, K713–K715, K717, K73, K74, K760, K762–K764, K768, K769, Z944 |
|  | | Diabetes without chronic complications | | E100, E101, E106, E108, E109, E110, E111, E116, E118, E119, E120, E121, E126, E128, E129, E130, E131, E136, E138, E139, E140, E141, E146, E148, E149 |
|  | | Diabetes with chronic complications | | E102–E105, E107, E112–E115, E117, E122–E125, E127, E132–E135, E137, E142–E145, E147 |
|  | | Hemiplegia or paraplegia | | G041, G114, G801, G802, G81, G82, G830–G834, G839 |
|  | | Renal disease | | I120, I131, N032–N037, N052–N057, N18, N19, N250, Z490–Z492, Z940, Z992 |
|  | | Any malignancy | | C00–C26, C30–C34, C37–C41, C43, C45–C58, C60–C76, C81–C85, C88, C90–C97 |
|  | | Moderate or severe liver disease | | I850, I859, I864, I982, K704, K711, K721, K729, K765–K767 |
|  | | Metastatic solid tumor | | C77–C80 |
|  | | AIDS/HIV | | B20–B22, B24 |
|  | | **Covariates for the bronchial asthma subgroup** * The following variables were defined from 365 days to 14 days before each time point. | | |
|  | | Systemic glucocorticoids | | H02AB (Oral use only. At least two prescriptions are required.) |
|  | | Category of inhaled drugs | | The categories of inhaled drugs were defined as follows: none, monotherapy (LABA or ICS), dual therapy (LABA and ICS), and triple therapy (LABA + ICS + long-acting muscarinic antagonists [LAMA]). (Priority: Triple therapy > Dual therapy > Monotherapy > None)  - ICS: R03BA, R03AK06, R03AK07, R03AK10, R03AK11, R03AK14, R03AL08, R03AL12 - LABA: R03AC12, R03AC13, R03AC18, R03AK06, R03AK07, R03AK10, R03AK11, R03AK14, R03AL08, R03AL12, R03CC11 (External use only), R03CC13 - Triple therapy: R03AL08, R03AL12 |
|  | | **Covariates for the atopic dermatitis subgroup** * The following variable were defined based on records from 365 days to 14 days before each time point. | | |
|  | | Topical glucocorticoids (≥very strong category) | | WHO-ATC code: D05AX52, D07AB02, D07AB11, D07AC01, D07AC06, D07AC08, D07AC09, D07AC11, D07AC13, D07AC19, D07AC10, D07AD01 |
|  | | Systemic glucocorticoids^†^ | | WHO-ATC code: H02AB (Oral use only.) |
|  | | Cyclosporin A^†^ | | WHO-ATC code: L04AD01 |
|  | | Phototherapy | | Category code: J054 |
| Abbrevidations: ICD-10, International Classification of Diseases, Tenth Revision; WHO-ATC code, Anatomical Therapeutic Chemical code by the World Health Organization.  † At least two prescriptions are required.  ‡ "YJ code" in the Japanese coding system | | | | |

| **Supplementary Table 2. Detailed diagnoses among patients with AAV in the nested case-control analysis** | | | | | | |
| --- | --- | --- | --- | --- | --- | --- |
|  | **anti-IL-5 (-)** | **anti-IL-5 (+)** | **anti-IL-4 (-)** | **anti-IL-4 (+)** | **anti-IgE (-)** | **anti-IgE (+)** |
| **Total AAV, *n*** | **734** | **14** | **741** | **7** | **744** | **4** |
| EGPA | 291 (39.6%) | 12 (85.7%) | 299 (40.4%) | 4 (57.1%) | 300 (40.3%) | 3 (75.0%) |
| GPA | 4 (0.5%) | 0 (0.0%) | 4 (0.5%) | 0 (0.0%) | 4 (0.5%) | 0 (0.0%) |
| MPA | 94 (12.8%) | 1 (7.1%) | 95 (12.8%) | 0 (0.0%) | 95 (12.8%) | 0 (0.0%) |
| Unspecified | 267 (36.4%) | 2 (14.3%) | 265 (35.8%) | 4 (57.1%) | 268 (36.0%) | 1 (25.0%) |
| IL: interleukin; EGPA: eosinophilic granulomatosis with polyangiitis; GPA: granulomatosis with polyangiitis; MPA: microscopic polyangiitis | | | | | | |

| **Supplementary Table 3. Effect of exposure drugs on the outcomes: BA patients** | | | | | | | | | | |
| --- | --- | --- | --- | --- | --- | --- | --- | --- | --- | --- |
|  |  | **NCC** | | | |  | **tCox** | | | |
| **Disease** | | **Case Ex/N** | **Control Ex/N** | **Crude** | **Adjusted** |  | **Ex Ev/PY** | **Control Ev/PY** | **Crude** | **Adjusted** |
| **Anti-IL-5 agent** | |  |  |  |  |  |  |  |  |  |
|  | **Composite** | 19/5690 | 22/34140 | **5.18 [2.81, 9.57]** | 1.22 [0.62, 2.42] |  | 16/836.1 | 5674/1574098.8 | **5.59 [3.42, 9.15]** | **2.00 [1.21, 3.31]** |
|  | **RA** | 12/1496 | 6/8976 | **12.00 [4.50, 31.97]** | **3.66 [1.17, 11.46]** |  | 12/1328.9 | 1484/1610376.4 | **10.30 [5.83, 18.18]** | **2.78 [1.55, 4.99]** |
|  | **SLE** | 7/378 | 3/2268 | **14.00 [3.62, 54.14]** | 3.13 [0.42, 23.26] |  | 6/1304.6 | 372/1622461.0 | **21.59 [9.43, 49.42]** | **3.95 [1.65, 9.44]** |
|  | **SSc** | 1/235 | 1/1410 | 6.00 [0.38, 95.93] | 100< [0.00, 100<] |  | 1/1391.7 | 234/1625228.2 | 5.53 [0.77, 39.62] | 1.94 [0.27, 13.94] |
|  | **DM/PM** | 1/419 | 1/2514 | 6.00 [0.38, 95.93] | 1.05 [0.06, 19.70] |  | 1/1377.5 | 418/1624641.6 | 2.87 [0.40, 20.55] | 0.69 [0.10, 4.98] |
|  | **SS** | 1/1290 | 8/7740 | 0.75 [0.09, 6.00] | 0.20 [0.02, 2.08] |  | 1/1354.2 | 1289/1614841.1 | 1.01 [0.14, 7.20] | 0.37 [0.05, 2.65] |
|  | **MCTD** | 0/85 | 0/510 | NA [NA, NA] | NA [NA, NA] |  | 0/1401.2 | 85/1625967.9 | **0.00 [0.00, 0.00]** | **0.00 [0.00, 0.00]** |
|  | **AAV** | 14/597 | 3/3582 | **28.00 [8.05, 97.43]** | 2.83 [0.67, 11.85] |  | 11/924.5 | 586/1623866.4 | **35.10 [19.07, 64.62]** | **5.23 [2.66, 10.30]** |
|  | **LVV** | 0/49 | 0/294 | NA [NA, NA] | NA [NA, NA] |  | 0/1399.7 | 49/1626769.6 | **0.00 [0.00, 0.00]** | **0.00 [0.00, 0.00]** |
|  | **PAN** | 1/33 | 0/198 | 100< [0.00, 100<] | 100< [0.00, 100<] |  | 1/1385.6 | 32/1626916.5 | **41.41 [5.39, 100<]** | **8.87 [1.12, 70.44]** |
|  | **PMR** | 1/231 | 3/1386 | 2.00 [0.21, 19.23] | 0.88 [0.05, 16.33] |  | 1/1399.4 | 230/1625865.8 | 5.62 [0.78, 40.27] | 1.79 [0.25, 12.90] |
|  | **Psoriasis** | 5/2318 | 10/13908 | **3.00 [1.03, 8.78]** | 1.28 [0.40, 4.12] |  | 5/1372.3 | 2313/1607790.6 | **2.58 [1.07, 6.20]** | 1.20 [0.50, 2.90] |
| **Anti-IL-4 agent** | |  |  |  |  |  |  |  |  |  |
|  | **Composite** | 19/5690 | 32/34140 | **3.56 [2.02, 6.29]** | 1.49 [0.79, 2.79] |  | 16/1689.8 | 5674/1573245.2 | **2.83 [1.73, 4.64]** | 1.36 [0.82, 2.24] |
|  | **RA** | 3/1496 | 10/8976 | 1.80 [0.50, 6.54] | 1.13 [0.24, 5.29] |  | 2/1843.8 | 1494/1609861.5 | 1.25 [0.31, 5.01] | 0.56 [0.14, 2.26] |
|  | **SLE** | 0/378 | 3/2268 | 0.00 [0.00, 100<] | 0.00 [0.00, 100<] |  | 0/1850.8 | 378/1621914.8 | **0.00 [0.00, 0.00]** | **0.00 [0.00, 0.00]** |
|  | **SSc** | 0/235 | 2/1410 | 0.00 [0.00, 100<] | 0.00 [0.00, 100<] |  | 0/1857.7 | 235/1624762.1 | **0.00 [0.00, 0.00]** | **0.00 [0.00, 0.00]** |
|  | **DM/PM** | 0/419 | 5/2514 | 0.00 [0.00, 100<] | 0.00 [0.00, 100<] |  | 0/1856.1 | 419/1624163.0 | **0.00 [0.00, 0.00]** | **0.00 [0.00, 0.00]** |
|  | **SS** | 5/1290 | 12/7740 | 2.50 [0.88, 7.10] | 2.94 [0.84, 10.30] |  | 5/1835.6 | 1285/1614359.7 | 3.93 [1.63, 9.48] | 2.35 [0.96, 5.78] |
|  | **MCTD** | 0/85 | 1/510 | 0.00 [0.00, 100<] | 0.00 [0.00, 100<] |  | 0/1859.3 | 85/1625509.8 | **0.00 [0.00, 0.00]** | **0.00 [0.00, 0.00]** |
|  | **AAV** | 7/597 | 7/3582 | **6.00 [2.11, 17.11]** | 1.58 [0.27, 9.35] |  | 6/1817.9 | 591/1622973.0 | **9.60 [4.22, 21.80]** | 2.10 [0.89, 4.99] |
|  | **LVV** | 0/49 | 0/294 | NA [NA, NA] | NA [NA, NA] |  | 0/1859.3 | 49/1626309.9 | **0.00 [0.00, 0.00]** | **0.00 [0.00, 0.00]** |
|  | **PAN** | 0/33 | 0/198 | NA [NA, NA] | NA [NA, NA] |  | 0/1859.3 | 33/1626442.7 | **0.00 [0.00, 0.00]** | **0.00 [0.00, 0.00]** |
|  | **PMR** | 0/231 | 1/1386 | 0.00 [0.00, 100<] | 0.00 [0.00, 100<] |  | 0/1859.3 | 231/1625405.8 | **0.00 [0.00, 0.00]** | **0.00 [0.00, 0.00]** |
|  | **Psoriasis** | 7/2318 | 17/13908 | **2.47 [1.03, 5.96]** | 1.20 [0.46, 3.18] |  | 6/1766.9 | 2312/1607396.1 | **2.48 [1.11, 5.54]** | 1.52 [0.68, 3.37] |
| **Anti-IgE agent** | |  |  |  |  |  |  |  |  |  |
|  | **Composite** | 8/5690 | 9/34140 | **5.33 [2.06, 13.82]** | 1.62 [0.57, 4.64] |  | 7/726.5 | 5683/1574208.4 | **2.73 [1.31, 5.70]** | 1.29 [0.61, 2.71] |
|  | **RA** | 1/1496 | 5/8976 | 1.20 [0.14, 10.27] | 0.54 [0.05, 6.25] |  | 1/780.6 | 1495/1610924.7 | 1.40 [0.20, 9.97] | 0.56 [0.08, 4.02] |
|  | **SLE** | 1/378 | 1/2268 | 6.00 [0.38, 95.93] | 17.74 [0.24, 100<] |  | 1/777.0 | 377/1622988.6 | 5.87 [0.83, 41.70] | 1.46 [0.20, 10.85] |
|  | **SSc** | 0/235 | 1/1410 | 0.00 [0.00, 100<] | 0.00 [0.00, 100<] |  | 0/794.6 | 235/1625825.2 | **0.00 [0.00, 0.00]** | **0.00 [0.00, 0.00]** |
|  | **DM/PM** | 1/419 | 1/2514 | 6.00 [0.38, 95.93] | 100< [0.00, 100<] |  | 0/793.3 | 419/1625225.8 | **0.00 [0.00, 0.00]** | **0.00 [0.00, 0.00]** |
|  | **SS** | 2/1290 | 5/7740 | 2.40 [0.47, 12.37] | 1.36 [0.20, 9.33] |  | 2/781.3 | 1288/1615413.9 | 3.35 [0.84, 13.35] | 1.66 [0.41, 6.71] |
|  | **MCTD** | 0/85 | 1/510 | 0.00 [0.00, 100<] | 0.00 [0.00, 100<] |  | 0/794.5 | 85/1626574.6 | 0.00 [0.00, 0.00] | 0.00 [0.00, 0.00] |
|  | **AAV** | 2/597 | 4/3582 | 3.00 [0.55, 16.38] | 0.38 [0.05, 2.66] |  | 2/789.8 | 595/1624001.1 | **7.18 [1.80, 28.72]** | 1.52 [0.35, 6.54] |
|  | **LVV** | 0/49 | 0/294 | NA [NA, NA] | NA [NA, NA] |  | 0/795.7 | 49/1627373.6 | **0.00 [0.00, 0.00]** | **0.00 [0.00, 0.00]** |
|  | **PAN** | 0/33 | 0/198 | NA [NA, NA] | NA [NA, NA] |  | 0/795.7 | 33/1627506.4 | **0.00 [0.00, 0.00]** | **0.00 [0.00, 0.00]** |
|  | **PMR** | 0/231 | 1/1386 | 0.00 [0.00, 100<] | 0.00 [0.00, 100<] |  | 0/795.7 | 231/1626469.5 | **0.00 [0.00, 0.00]** | **0.00 [0.00, 0.00]** |
|  | **Psoriasis** | 2/2318 | 8/13908 | 1.50 [0.32, 7.06] | 0.65 [0.13, 3.31] |  | 2/774.3 | 2316/1608388.6 | 1.78 [0.45, 7.11] | 1.04 [0.26, 4.21] |
| The effect of the exposure drug is presented as the odds ratio [95% CI] for the nested case-control analysis and the hazard ratio [95% CI] for the time-dependent Cox analysis. Results with a 95% CI that does not include 1.0 are indicated in bold. Abbreviations: Ex, Exposure; Ev, Event; NA, Not applicable; NCC, Nested case-control analysis; PY, Patient-year; tCox, Time-dependent Cox analysis. | | | | | | | | | | |

| **Supplementary Table 4. Effect of exposure drugs on the outcomes: AD patients** | | | | | | | | | | |
| --- | --- | --- | --- | --- | --- | --- | --- | --- | --- | --- |
|  |  | **NCC** | | | |  | **tCox** | | | |
| **Disease** | | **Case Ex/N** | **Control Ex/N** | **Crude** | **Adjusted** |  | **Ex Ev/PY** | **Control Ev/PY** | **Crude** | **Adj** |
| **Anti-IL-5 agent** | |  |  |  |  |  |  |  |  |  |
|  | **Composite** | 1/5851 | 1/35106 | 6.00 [0.38, 95.93] | 4.32 [0.25, 75.98] |  | 1/15.1 | 5850/962743.9 | 13.45 [2.10, 86.23] | 4.24 [0.67, 27.02] |
|  | **RA** | 0/597 | 0/3582 | NA [NA, NA] | NA [NA, NA] |  | 0/35.5 | 597/1007986.4 | **0.00 [0.00, 0.00]** | **0.00 [0.00, 0.00]** |
|  | **SLE** | 1/205 | 0/1230 | 100< [0.00, 100<] | 100< [0.00, 100<] |  | 1/36.3 | 204/1010996.5 | 100< [28.92, 100<] | 24.59 [3.13, 100<] |
|  | **SSc** | 0/120 | 0/720 | NA [NA, NA] | NA [NA, NA] |  | 0/41.3 | 120/1012355.8 | **0.00 [0.00, 0.00]** | **0.00 [0.00, 0.00]** |
|  | **DM/PM** | 0/201 | 0/1206 | NA [NA, NA] | NA [NA, NA] |  | 0/40.4 | 201/1012031.4 | **0.00 [0.00, 0.00]** | **0.00 [0.00, 0.00]** |
|  | **SS** | 1/555 | 0/3330 | 100< [0.00, 100<] | 100< [0.00, 100<] |  | 1/34.8 | 554/1007619.1 | 59.48 [8.67, 100<] | 11.65 [1.53, 89.00] |
|  | **MCTD** | 0/50 | 0/300 | NA [NA, NA] | NA [NA, NA] |  | 0/41.3 | 50/1013004.9 | **0.00 [0.00, 0.00]** | **0.00 [0.00, 0.00]** |
|  | **AAV** | 0/128 | 0/768 | NA [NA, NA] | NA [NA, NA] |  | 0/25.0 | 128/1012835.4 | **0.00 [0.00, 0.00]** | **0.00 [0.00, 0.00]** |
|  | **LVV** | 0/24 | 0/144 | NA [NA, NA] | NA [NA, NA] |  | 0/38.9 | 24/1013473.7 | **0.00 [0.00, 0.00]** | **0.00 [0.00, 0.00]** |
|  | **PAN** | 0/15 | 0/90 | NA [NA, NA] | NA [NA, NA] |  | 0/41.3 | 15/1013464.3 | **0.00 [0.00, 0.00]** | **0.00 [0.00, 0.00]** |
|  | **PMR** | 0/76 | 0/456 | NA [NA, NA] | NA [NA, NA] |  | 0/36.8 | 76/1013274.4 | **0.00 [0.00, 0.00]** | **0.00 [0.00, 0.00]** |
|  | **Psoriasis** | 1/4666 | 0/27996 | 100< [0.00, 100<] | 100< [0.00, 100<] |  | 0/30.7 | 4666/975441.0 | **0.00 [0.00, 0.00]** | **0.00 [0.00, 0.00]** |
| **Anti-IL-4 agent** | |  |  |  |  |  |  |  |  |  |
|  | **Composite** | 35/5851 | 239/35106 | 0.88 [0.61, 1.25] | **0.50 [0.34, 0.74]** |  | 32/6620.5 | 5819/956138.5 | 1.05 [0.74, 1.48] | **0.64 [0.45, 0.91]** |
|  | **RA** | 7/597 | 26/3582 | 1.62 [0.70, 3.72] | 0.95 [0.33, 2.73] |  | 7/7443.0 | 590/1000578.9 | 1.76 [0.83, 3.74] | 1.20 [0.57, 2.56] |
|  | **SLE** | 1/205 | 11/1230 | 0.55 [0.07, 4.23] | 1.59 [0.16, 16.18] |  | 1/7470.0 | 204/1003562.8 | 1.05 [0.14, 7.62] | 0.59 [0.08, 4.50] |
|  | **SSc** | 1/120 | 5/720 | 1.20 [0.14, 10.27] | 4.40 [0.22, 86.23] |  | 1/7465.7 | 119/1004931.5 | 1.46 [0.20, 10.62] | 1.25 [0.17, 9.16] |
|  | **DM/PM** | 0/201 | 5/1206 | 0.00 [0.00, 100<] | 0.00 [0.00, 100<] |  | 0/7457.3 | 201/1004614.4 | **0.00 [0.00, 0.00]** | **0.00 [0.00, 0.00]** |
|  | **SS** | 5/555 | 28/3330 | 1.07 [0.41, 2.81] | 0.71 [0.21, 2.40] |  | 5/7371.0 | 550/1000282.9 | 1.41 [0.58, 3.41] | 1.04 [0.44, 2.48] |
|  | **MCTD** | 0/50 | 1/300 | 0.00 [0.00, 100<] | 0.00 [0.00, 100<] |  | 0/7467.3 | 50/1005579.0 | **0.00 [0.00, 0.00]** | **0.00 [0.00, 0.00]** |
|  | **AAV** | 0/128 | 1/768 | 0.00 [0.00, 100<] | 0.00 [0.00, 100<] |  | 0/7466.1 | 128/1005394.4 | **0.00 [0.00, 0.00]** | **0.00 [0.00, 0.00]** |
|  | **LVV** | 1/24 | 2/144 | 3.00 [0.27, 33.09] | 3.35 [0.04, 100<] |  | 1/7470.6 | 23/1006042.0 | 4.86 [0.60, 39.04] | 3.23 [0.38, 27.37] |
|  | **PAN** | 0/15 | 1/90 | 0.00 [0.00, 100<] | 0.00 [0.00, 100<] |  | 0/7471.7 | 15/1006033.9 | **0.00 [0.00, 0.00]** | **0.00 [0.00, 0.00]** |
|  | **PMR** | 0/76 | 3/456 | 0.00 [0.00, 100<] | 0.00 [0.00, 100<] |  | 0/7462.8 | 76/1005848.5 | **0.00 [0.00, 0.00]** | **0.00 [0.00, 0.00]** |
|  | **Psoriasis** | 30/4666 | 153/27996 | 1.18 [0.80, 1.74] | **0.54 [0.35, 0.85]** |  | 27/6734.2 | 4639/968737.4 | 1.16 [0.79, 1.69] | **0.68 [0.47, 1.00]** |
| **Anti-IgE agent** | |  |  |  |  |  |  |  |  |  |
|  | **Composite** | 9/5851 | 9/35106 | **6.00 [2.38, 15.12]** | 2.07 [0.70, 6.13] |  | 9/343.8 | 5842/962415.2 | **4.75 [2.47, 9.14]** | 1.85 [0.96, 3.59] |
|  | **RA** | 1/597 | 0/3582 | 100< [0.00, 100<] | 100< [0.00, 100<] |  | 1/397.8 | 596/1007624.0 | 4.28 [0.61, 30.15] | 1.07 [0.16, 7.31] |
|  | **SLE** | 0/205 | 0/1230 | NA [NA, NA] | NA [NA, NA] |  | 0/400.2 | 205/1010632.6 | **0.00 [0.00, 0.00]** | **0.00 [0.00, 0.00]** |
|  | **SSc** | 1/120 | 0/720 | 100< [0.00, 100<] | 100< [0.00, 100<] |  | 1/401.4 | 119/1011995.8 | **23.00 [3.18, 100<]** | **8.07 [1.23, 53.07]** |
|  | **DM/PM** | 0/201 | 0/1206 | NA [NA, NA] | NA [NA, NA] |  | 0/399.5 | 201/1011672.2 | **0.00 [0.00, 0.00]** | **0.00 [0.00, 0.00]** |
|  | **SS** | 0/555 | 1/3330 | 0.00 [0.00, 100<] | 0.00 [0.00, 100<] |  | 0/386.4 | 555/1007267.5 | **0.00 [0.00, 0.00]** | **0.00 [0.00, 0.00]** |
|  | **MCTD** | 0/50 | 0/300 | NA [NA, NA] | NA [NA, NA] |  | 0/402.6 | 50/1012643.6 | **0.00 [0.00, 0.00]** | **0.00 [0.00, 0.00]** |
|  | **AAV** | 1/128 | 0/768 | 100< [0.00, 100<] | 100< [0.00, 100<] |  | 1/400.8 | 127/1012459.6 | **22.10 [3.08, 100<]** | 6.66 [0.91, 49.02] |
|  | **LVV** | 1/24 | 0/144 | 100< [0.00, 100<] | 100< [0.00, 100<] |  | 1/402.0 | 23/1013110.6 | **99.83 [12.97, 100<]** | **39.67 [3.90, 100<]** |
|  | **PAN** | 0/15 | 0/90 | NA [NA, NA] | NA [NA, NA] |  | 0/402.6 | 15/1013103.0 | **0.00 [0.00, 0.00]** | **0.00 [0.00, 0.00]** |
|  | **PMR** | 0/76 | 0/456 | NA [NA, NA] | NA [NA, NA] |  | 0/401.4 | 76/1012909.8 | **0.00 [0.00, 0.00]** | **0.00 [0.00, 0.00]** |
|  | **Psoriasis** | 6/4666 | 8/27996 | **4.50 [1.56, 12.97]** | 1.04 [0.31, 3.45] |  | 6/364.3 | 4660/975107.4 | **3.86 [1.74, 8.54]** | 1.57 [0.70, 3.51] |
| The effect of the exposure drug is presented as the odds ratio [95% CI] for the nested case-control analysis and the hazard ratio [95% CI] for the time-dependent Cox analysis. Results with a 95% CI that does not include 1.0 are indicated in bold. Abbreviations: Ex, Exposure; Ev, Event; NA, Not applicable; NCC, Nested case-control analysis; PY, Patient-year; tCox, Time-dependent Cox analysis. | | | | | | | | | | |

| **Supplementary Table 5. Effect of exposure drugs on the outcomes: sensitivity analysis (washout period =42 days)** | | | | | | | | | | |
| --- | --- | --- | --- | --- | --- | --- | --- | --- | --- | --- |
|  |  | **NCC** | | | |  | **tCox** | | | |
| **Disease** | | **Case Ex/N** | **Control Ex/N** | **Crude** | **Adjusted** |  | **Ex Ev/PY** | **Control Ev/PY** | **Crude** | **Adjusted** |
| **Anti-IL-5 agent** | |  |  |  |  |  |  |  |  |  |
|  | **Composite** | 16/11989 | 25/71934 | **3.84 [2.05, 7.19]** | **2.56 [1.31, 5.02]** |  | 16/815.9 | 11973/2662527.2 | **5.11 [3.13, 8.35]** | **2.98 [1.81, 4.91]** |
|  | **RA** | 12/2195 | 6/13170 | **12.00 [4.50, 31.97]** | **9.29 [2.93, 29.45]** |  | 12/1317.2 | 2183/2747456.2 | **12.24 [6.94, 21.61]** | **5.23 [2.92, 9.35]** |
|  | **SLE** | 7/602 | 1/3612 | **42.00 [5.17, 100<]** | **33.66 [2.88, 100<]** |  | 6/1298.4 | 596/2763382.9 | **26.08 [11.53, 59.00]** | **10.99 [4.72, 25.58]** |
|  | **SSc** | 1/379 | 3/2274 | 2.00 [0.21, 19.23] | 0.73 [0.06, 9.19] |  | 1/1383.7 | 378/2767607.1 | 6.10 [0.86, 43.56] | 2.67 [0.36, 19.56] |
|  | **DM/PM** | 1/653 | 1/3918 | 6.00 [0.38, 95.93] | 1.86 [0.08, 41.21] |  | 1/1369.1 | 652/2766673.4 | 3.45 [0.48, 24.69] | 1.67 [0.23, 11.95] |
|  | **SS** | 2/1950 | 7/11700 | 1.71 [0.36, 8.25] | 0.82 [0.13, 5.14] |  | 2/1341.1 | 1948/2751687.6 | 2.34 [0.58, 9.37] | 1.03 [0.26, 4.13] |
|  | **MCTD** | 0/143 | 0/858 | NA [NA, NA] | NA [NA, NA] |  | 0/1393.0 | 143/2769090.4 | **0.00 [0.00, 0.00]** | **0.00 [0.00, 0.00]** |
|  | **AAV** | 9/748 | 1/4488 | **54.00 [6.84, 100<]** | **18.90 [2.14, 100<]** |  | 9/909.6 | 739/2766687.8 | **40.47 [20.76, 78.90]** | **12.20 [5.82, 25.55]** |
|  | **LVV** | 0/75 | 0/450 | NA [NA, NA] | NA [NA, NA] |  | 0/1390.7 | 75/2770414.6 | **0.00 [0.00, 0.00]** | **0.00 [0.00, 0.00]** |
|  | **PAN** | 1/51 | 0/306 | 100< [0.00, 100<] | 100< [0.00, 100<] |  | 1/1372.6 | 50/2770553.4 | **46.59 [6.30, 100<]** | **24.73 [3.14, 100<]** |
|  | **PMR** | 1/325 | 2/1950 | 3.00 [0.27, 33.09] | 0.57 [0.03, 9.47] |  | 1/1386.9 | 324/2769234.6 | 6.67 [0.94, 47.62] | 2.59 [0.37, 18.35] |
|  | **Psoriasis** | 4/7214 | 21/43284 | 1.14 [0.39, 3.33] | 1.33 [0.44, 4.02] |  | 4/1359.3 | 7210/2711303.1 | 1.37 [0.52, 3.65] | 1.27 [0.47, 3.38] |
| **Anti-IL-4 agent** | |  |  |  |  |  |  |  |  |  |
|  | **Composite** | 48/11989 | 234/71934 | 1.23 [0.90, 1.68] | 0.73 [0.53, 1.01] |  | 42/7955.1 | 11947/2655388.0 | **1.40 [1.03, 1.89]** | 0.94 [0.69, 1.27] |
|  | **RA** | 9/2195 | 38/13170 | 1.43 [0.69, 2.98] | 2.11 [0.81, 5.53] |  | 9/8868.9 | 2186/2739904.5 | 1.37 [0.71, 2.64] | 1.30 [0.67, 2.52] |
|  | **SLE** | 1/602 | 14/3612 | 0.43 [0.06, 3.26] | 0.28 [0.03, 2.57] |  | 1/8900.9 | 601/2755780.4 | 0.63 [0.09, 4.49] | 0.45 [0.06, 3.24] |
|  | **SSc** | 1/379 | 6/2274 | 1.00 [0.12, 8.31] | 0.28 [0.02, 5.38] |  | 1/8903.3 | 378/2760087.5 | 0.95 [0.13, 6.83] | 0.95 [0.13, 6.86] |
|  | **DM/PM** | 0/653 | 21/3918 | 0.00 [0.00, 100<] | 0.00 [0.00, 100<] |  | 0/8895.7 | 653/2759146.8 | **0.00 [0.00, 0.00]** | **0.00 [0.00, 0.00]** |
|  | **SS** | 8/1950 | 40/11700 | 1.20 [0.56, 2.58] | 1.13 [0.49, 2.61] |  | 8/8795.1 | 1942/2744233.6 | 1.45 [0.72, 2.90] | 1.42 [0.71, 2.84] |
|  | **MCTD** | 0/143 | 2/858 | 0.00 [0.00, 100<] | 0.00 [0.00, 100<] |  | 0/8907.2 | 143/2761576.2 | **0.00 [0.00, 0.00]** | **0.00 [0.00, 0.00]** |
|  | **AAV** | 6/748 | 17/4488 | 2.15 [0.84, 5.51] | 1.23 [0.42, 3.60] |  | 6/8863.7 | 742/2758733.7 | 2.72 [1.21, 6.12] | 2.20 [0.95, 5.07] |
|  | **LVV** | 1/75 | 1/450 | 6.00 [0.38, 95.93] | 5.62 [0.14, 100<] |  | 1/8910.3 | 74/2762895.0 | 4.58 [0.61, 34.27] | 4.27 [0.57, 31.95] |
|  | **PAN** | 0/51 | 0/306 | NA [NA, NA] | NA [NA, NA] |  | 0/8911.4 | 51/2763014.5 | **0.00 [0.00, 0.00]** | **0.00 [0.00, 0.00]** |
|  | **PMR** | 0/325 | 11/1950 | 0.00 [0.00, 100<] | 0.00 [0.00, 100<] |  | 0/8902.7 | 325/2761718.8 | **0.00 [0.00, 0.00]** | **0.00 [0.00, 0.00]** |
|  | **Psoriasis** | 34/7214 | 108/43284 | **1.90 [1.29, 2.79]** | 0.86 [0.57, 1.29] |  | 28/8137.4 | 7186/2704525.0 | **1.63 [1.12, 2.36]** | 0.87 [0.60, 1.26] |
| **Anti-IgE agent** | |  |  |  |  |  |  |  |  |  |
|  | **Composite** | 28/11989 | 31/71934 | **5.42 [3.25, 9.03]** | **2.99 [1.74, 5.13]** |  | 28/1818.3 | 11961/2661524.8 | **3.55 [2.45, 5.15]** | **2.50 [1.72, 3.63]** |
|  | **RA** | 3/2195 | 3/13170 | **6.00 [1.21, 29.73]** | 3.43 [0.36, 32.37] |  | 3/1966.0 | 2192/2746807.4 | 1.91 [0.62, 5.92] | 1.08 [0.35, 3.36] |
|  | **SLE** | 1/602 | 0/3612 | 100< [0.00, 100<] | 100< [0.00, 100<] |  | 1/1968.1 | 601/2762713.2 | 2.46 [0.35, 17.45] | 1.27 [0.17, 9.44] |
|  | **SSc** | 2/379 | 1/2274 | **12.00 [1.09, 100<]** | 16.04 [0.82, 100<] |  | 2/1991.7 | 377/2766999.1 | **7.70 [1.92, 30.96]** | **4.60 [1.13, 18.76]** |
|  | **DM/PM** | 1/653 | 1/3918 | 6.00 [0.38, 95.93] | 16.86 [0.75, 100<] |  | 1/1987.0 | 652/2766055.5 | 2.24 [0.31, 15.90] | 1.42 [0.20, 10.16] |
|  | **SS** | 4/1950 | 10/11700 | 2.40 [0.75, 7.65] | 1.49 [0.42, 5.38] |  | 4/1955.6 | 1946/2751073.1 | **2.94 [1.10, 7.84]** | 1.62 [0.60, 4.34] |
|  | **MCTD** | 1/143 | 0/858 | 100< [0.00, 100<] | 100< [0.00, 100<] |  | 1/1992.9 | 142/2768490.4 | **9.95 [1.39, 71.19]** | 5.80 [0.81, 41.58] |
|  | **AAV** | 4/748 | 2/4488 | **12.00 [2.20, 65.52]** | 5.32 [0.84, 33.72] |  | 4/1978.0 | 744/2765619.4 | **7.77 [2.91, 20.75]** | **3.93 [1.38, 11.24]** |
|  | **LVV** | 1/75 | 1/450 | 6.00 [0.38, 95.93] | 6.81 [0.36, 100<] |  | 1/1993.8 | 74/2769811.5 | **18.40 [2.54, 100<]** | **14.39 [1.94, 100<]** |
|  | **PAN** | 0/51 | 0/306 | NA [NA, NA] | NA [NA, NA] |  | 0/1994.5 | 51/2769931.4 | **0.00 [0.00, 0.00]** | **0.00 [0.00, 0.00]** |
|  | **PMR** | 0/325 | 0/1950 | NA [NA, NA] | NA [NA, NA] |  | 0/1993.4 | 325/2768628.1 | **0.00 [0.00, 0.00]** | **0.00 [0.00, 0.00]** |
|  | **Psoriasis** | 17/7214 | 23/43284 | **4.44 [2.37, 8.30]** | **2.69 [1.35, 5.37]** |  | 17/1907.1 | 7197/2710755.3 | **3.53 [2.19, 5.68]** | **2.76 [1.71, 4.45]** |
| The effect of the exposure drug is presented as the odds ratio [95% CI] for the nested case-control analysis and the hazard ratio [95% CI] for the time-dependent Cox analysis. Results with a 95% CI that does not include 1.0 are indicated in bold. Abbreviations: Ex, Exposure; Ev, Event; NA, Not applicable; NCC, Nested case-control analysis; PY, Patient-year; tCox, Time-dependent Cox analysis. | | | | | | | | | | |

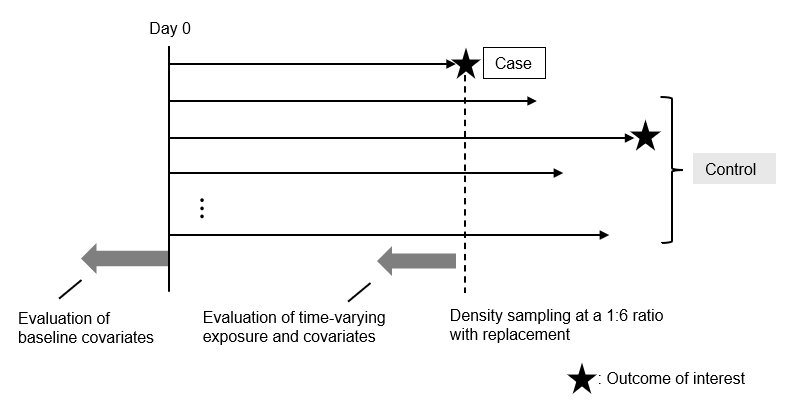
**Supplementary Figure 1. Graphical representation of the nested case-control analysis**

In the nested case-control analysis, control patients were randomly selected using density sampling at a 1:6 ratio with replacement, based on the number of days from the cohort entry date (day 0). Time-varying variables were assessed based on the case date.
